# Randomized dose-response trial of n-3 fatty acids in hormone receptor negative breast cancer survivors—impact on breast adipose oxylipin and DNA methylation patterns

**DOI:** 10.1101/2024.09.16.24313691

**Authors:** David E. Frankhouser, Todd DeWess, Isabel F. Snodgrass, Rachel M. Cole, Sarah Steck, Danielle Thomas, Chidimma Kalu, Martha A. Belury, Steven K. Clinton, John W. Newman, Lisa D. Yee

## Abstract

**Background:** Increasing evidence suggests the unique susceptibility of estrogen receptor and progesterone receptor negative (ERPR-) breast cancer to dietary fat amount and type. Dietary n-3 polyunsaturated fatty acids (PUFAs), such as docosahexaenoic acid (DHA) and eicosapentaenoic acid (EPA), may modulate breast adipose fatty acid profiles and downstream bioactive metabolites to counteract pro-inflammatory, pro-carcinogenic signaling in the mammary microenvironment.

**Objective:** To determine effects of ∼1 to 5 g/d EPA+DHA over 12 months on breast adipose fatty acid and oxylipin profiles in women with ERPR(−) breast cancer, a high-risk molecular subtype.

**Methods:** We conducted a 12-month randomized controlled, double-blind clinical trial of ∼5g/d vs ∼1g/d DHA+EPA supplementation in women within 5 years of completing standard therapy for ERPR(−) breast cancer Stages 0-III. Blood and breast adipose tissue specimens were collected every 3 months for biomarker analyses including fatty acids by gas chromatography, oxylipins by LC-MS/MS, and DNA methylation by reduced-representation bisulfite sequencing (RRBS).

**Results:** A total of 51 participants completed the 12-month intervention. Study treatments were generally well-tolerated. While both doses increased n-3 PUFAs from baseline in breast adipose, erythrocytes, and plasma, the 5g/d supplement was more potent (n =51, p <0.001). The 5g/d dose also reduced plasma triglycerides from baseline (p =0.008). Breast adipose oxylipins at 0, 6, and 12 months showed dose-dependent increases in unesterified and esterified DHA and EPA metabolites (n =28). Distinct DNA methylation patterns in adipose tissue after 12 months were identified, with effects unique to the 5g/d dose group (n =17).

**Conclusions:** Over the course of 1 year, EPA+DHA dose-dependently increased concentrations of these fatty acids and their derivative oxylipin metabolites, producing differential DNA methylation profiles of gene promoters involved in metabolism-related pathways critical to ERPR(−) breast cancer development and progression. These data provide evidence of both metabolic and epigenetic effects of n-3 PUFAs in breast adipose tissue, elucidating novel mechanisms of action for high-dose EPA+DHA-mediated prevention of ERPR(−) breast cancer.

Clinicaltrials.gov identifier NCT02295059

## Introduction

Dietary fat composition may differentially affect susceptibility to specific types of breast cancer. Of longstanding interest for breast cancer prevention are eicosapentaenoic acid (EPA) and docosahexaenoic acid (DHA), the primary long chain n-3 PUFAs found in the oils of cold water fish and shellfish, and certain emerging specialty foods.^1^ Our pre-clinical studies demonstrated that diets enriched in fish oil decrease the incidence and multiplicity of ERPR(−)HER2/neu(+) mammary tumors in HER2/neu transgenic mice.^2^ Similarly, Turbitt et al. showed that fish oil vs corn oil diets inhibited mammary tumorigenesis in HER2/neu transgenic mice but not PyMT mice which develop ERPR(+)HER2(+) tumors.^3^ These findings raise the possibility that human breast cancers with the poor prognostic features of ERPR(−)±HER-2/neu overexpression might be uniquely responsive to EPA and DHA.

In prior clinical trials of women at high risk of breast cancer, we showed significantly increased breast adipose EPA and DHA following EPA+DHA capsule supplementation or fish consumption within 3 months.^4,5^ Emerging data indicate that metabolites of EPA and DHA, such as the oxylipins formed by the enzymatic and non-enzymatic oxidation of PUFAs, may mediate at least in part the physiologic effects of dietary n-3 PUFAs. Studies of n-3 PUFA supplementation in healthy volunteers have demonstrated dose-dependent increases in circulating EPA– and DHA-derived oxylipins with pro-resolving properties.^6,7^ In patients with morbid obesity treated with EPA+DHA supplements prior to bariatric surgery, analysis of visceral and subcutaneous fat showed increased EPA– and DHA-derived anti-inflammatory oxylipins such as 18-hydroxyeicosapentaenoic acid (HEPE) and 17-hydroxydocosoahexaenoic acid (HDoHE), respectively.^8^ However, how such dietary exposures influence mammary adipose tissue fatty acid and oxylipin composition is unknown. Enriching breast adipose tissue with anti-inflammatory/antiproliferative metabolites could suppress pro-carcinogenic processes in an at-risk mammary microenvironment.^9^

Here, we conducted a 12-month randomized controlled double-blind trial of EPA+DHA supplementation at either ∼5g/d or ∼1g/d in women within 5 years of completing standard therapy for triple negative or ERPR(−) HER2(+) breast cancer. The primary objective was to determine the effects of n-3 PUFA dose and duration on breast adipose fatty acid and oxylipin profiles in women with history of high risk ERPR(−) breast cancer. Secondary objectives included assessment of adipose tissue changes in DNA methylation (DNAm) as a modulating factor of gene expression and metabolism.

## Methods

### Study Oversight

This was a multi-institutional randomized controlled double-blind trial of high-dose EPA+DHA supplementation versus low-dose EPA+DHA supplementation approved by the Institutional Review Boards of The Ohio State University (OSU) and City of Hope (COH). The trial was conducted in accordance with institutional guidelines. Enrollment opened 9/2014 at OSU and 6/2017 at COH, with completion of accrual 11/2018 and last study visit 11/2019. Study participants were informed of the investigational nature of the study and provided written informed consent.

### Patient Eligibility Criteria

Eligible participants were women >18 years with Stage 0 to III ERPR(−) breast cancer who had completed standard of care treatment ≤5 years, with completion of chemotherapy >6 months, trastuzumab >3 months, and radiation therapy ≥2 months, as applicable. Ineligibility criteria included history of radiation to both breasts or bilateral mastectomies, chronic use of n-3 fatty acid concentrates or capsules within 3 months prior to enrollment, dietary consumption >2 fish servings/week, pregnancy/lactation, known sensitivity or allergy to fish, use of anticoagulant medications or standing regimen of full dose aspirin (≥ 325 mg/day), NSAIDs or NSAID-containing products.

### Study Intervention

Following signed informed consent, study participants were scheduled for the baseline visit and randomized to ∼5 g EPA+DHA/d or ∼1 g EPA+DHA/d. Women were instructed to maintain their baseline diet. The two EPA+DHA dose levels were selected from our prior n-3 PUFA dose-defining trial in women at high risk for breast cancer (∼5.03 g and ∼0.84 g EPA+DHA/d).^5^ Only the pharmacist and biostatistician were aware of randomization assignment. Women were advised to take 5 capsules/day in divided doses for 12 months. Study capsules for high-dose (MNSG-440W) and matching low-dose (MNSG-440PW) were manufactured by Marine Ingredients (Mt. Bethel, PA) to specifications of EPA+DHA as triglyceride of 1000 mg/capsule and 166.67 mg/capsule, respectively. Two batches for each dose level were utilized over the course of the trial, providing high and low mean EPA+DHA doses at 5.23 g/d and 0.91 g/d, respectively. EPA+DHA in the lower dose capsules were combined with a mix of saturated, monounsaturated, and polyunsaturated fatty acids to match the amount of fat provided by capsules in each arm (**Supplementary Table S1**). Both study agents contained a proprietary natural antioxidant blend of tocopherols, rosemary extract, and ascorbyl palmitate with natural lemon flavor with shell ingredients of gelatin, glycerin, purified water, and white color coating to appear identical for blinding purposes.

Participants were instructed to complete daily logs of oral doses of study capsules taken in addition to changes in medication and symptoms or adverse events. Daily logs and pill counts were reviewed at study visits every 3 months. Anthropometric measurements (e.g., body weight, height, BMI, waist circumference/hip circumference (WHR) were taken at 0, 3, 6, 9, and 12 months. Research biospecimens (breast adipose tissue fine needle aspiration and fasting blood samples) were collected at each study visit and processed and stored at –80°C for future batch analysis. Blood samples were also processed by clinical laboratories at each institution for alanine aminotransferase (ALT), a standard clinical lipid panel (i.e. total cholesterol, HDL-C, LDL-C, triglycerides), insulin, and glucose at 0, 6, and 12 months, along with platelet function assay at 0 and 12 months on a PFA-100 System (Siemens, Marburg, Germany).

### Fatty acid analyses

Fatty acids were quantified as methyl esters by gas chromatography (GC) with flame ionization detection. Briefly, total lipids were extracted from plasma, erythrocyte membrane, and breast adipose samples with 2:1 v/v chloroform/methanol and a 0.88% KCLwash.^10^ The fatty acid methyl esters were then prepared using 5% hydrochloric acid in methanol at 100°C.^11^ Erythrocyte lipids were extracted and methyl esters were prepared using borontrifluoride in methanol.^12^ Analysis of fatty acid methyl esters was completed by GC with column and conditions as previously described.^13^ Retention times were compared to standards (Matreya, LLC, Pleasant Gap, PA, Supelco®, Bellefonte, PA, and Nu-Check Prep Inc, Elysian, MN) and fatty acids are reported as weight percent of total identified.

### Esterified and non-esterified lipid mediator analysis

Lipid mediators were isolated in the presence of isotopically labeled standards, separated by ultraperformance liquid chromatography (UPLC), detected by electrospray ionization tandem mass spectrometry (MS/MS) and quantified against authentic calibration standards.

#### Sample extraction and ester hydrolysis

Non-esterified oxylipins were measured by UPLC-MS/MS after protein precipitation in the presence of isotopically labeled analytical surrogates as previously described.^14^ Briefly, ∼50mg of tissue were enriched with deuterated oxylipins, and homogenized on a vertical ball mill, GenoGrinder High Throughput Sample Homogenizer SPEX (RRID:SCR_019966), in 1:1 methanol/acetonitrile in the presence of a suite of deuterated oxylipin free acids and 1-cyclohexyl uriedo-3-dodecanoic acid (CUDA; Cayman Chemical; Ann Arbor, MI). Homogenates were centrifuged for 3min at 2.3g and 4°C, chilled at –20°C for 15 min and filtered with 0.2µm PVDF 96-well plates (Agilent Technologies; Santa Clara, CA). Samples were stored at –20°C until analysis within 48h.

Esterified oxylipins were measured by UPLC-MS/MS after tissue extraction with 8:10:11 isopropanol/cyclohexane/ammonium acetate, followed by alkaline hydrolysis and isolation by solid phase extraction using modifications of previously reported methods.^14^ Briefly, tissues were subsampled on dry ice with ∼50mg tissue mixed with 5 µL 0.2 mg/mL butylated hydroxy toluene/EDTA and homogenized in 410 µL isopropanol. Samples were then mixed with 520 µL cyclohexane followed by 570 µL of 0.1M ammonium acetate and centrifuged at 4°C. The supernatant was isolated, the homogenate was extracted with an additional 520 µL cyclohexane, and the supernatants combined. Solvents were removed by vacuum evaporation and the residue dissolved in 100µL 1:1 methanol/toluene (v/v). The lipid extracts were then enriched with a suite of deuterated oxylipin free acids, incubated with 100 µL 0.5M sodium methoxide for 50min at 50 °C, mixed with 100 µL water, and returned to 50 °C for 50min. Samples were neutralized with 10 µL 20% glacial acetic acid, diluted with 0.5 mL 0.1% acetic acid/5% MeOH, and oxylipins were trapped on a 10mg Oasis HLB solid phase extraction 96 well plates (Waters Corp, Milford Mass). After washing with 2 mL 0.1% acetic acid / 30%MeOH, analytes were eluted with 250 µL methanol followed by 1 mL ethyl acetate and collected into plate wells containing 10µL pf 20% glycerol in methanol. Solvents were removed by vacuum evaporation and residues reconstituted in 125µL of 1:1 MeOH/acetonitrile containing 100 nM CUDA, the sample was chilled and filtered at 0.2µm as above. Samples were stored at –20°C until analysis within 48h.

#### Lipid mediator quantification

Analytes were chromatographically separated on a Shimadzu Nexera X2 UHPLC (Shimadzu, Columbia, MD) using a 2.1 x 150mm Acquity BEH_C18_ column (Waters Corp) and quantified on an AB Sciex 6500 QTRAP (Sciex; Redwood City, CA) using electrospray ionization as previously described.^15^ The majority of oxylipins were quantified using isotope dilution and internal standard ratio-response methodologies against a minimum 7pt calibration curve bracketing reported concentrations. F2-isoprostanes (F2isoPs) were detected as a complex cluster of analytes with the same mass transition as PGF2a and surrounding the PGF2a peak. F2-IsoP concentrations were estimated using the response of prostaglandin F2a (353.3 > 193.2 m/z) as previously described.^16^ Due to sample heterogeneity, replicate precision was assessed and determined to be acceptable. Low recoveries of deuterated 13-oxo-ODE with the sodium methoxide ester hydrolysis were noted, and ketones were excluded from the esterified data set. Results are expressed in pmol/g tissue (i.e., nM) except for a subset of analytes, including PUFAs and 6 alpha-linolenic acid (ALA)-derived oxylipins for which commercially available standards were not available. For these 6 oxylipins, peaks were identified against non-quantitative authentic standards. The UPLC-MS/MS results for these metabolites are reported as relative abundance across all measured samples (i.e., the sum of each metabolite across all samples is set to 100%).

Storage time (5.5 ± 0.8 y prior to analysis) minimally affected a subset of metabolites. In the esterified oxylipin pool, of the 83 metabolites detected in >90% of samples, 4 EPA-derived alcohols and one EPA-derived diol significantly decreased with storage time. However, this explained only 3 ± 3% of the variance in these metabolites. In addition, 2 non-vicinal diols, 3 arachidonic acid (AA)-derived diols, 1 ALA-derived diol, 2 N-acylethanolamides and arachidonoyl serine increased with storage time and explained between 2 and 12% of the variance in these compounds. Therefore, while changes in metabolite concentrations were detected with storage time, these changes were small. Regardless, storage time was included as a fixed effect in least square regression models.

### Statistical analyses

The sample size (n=40/group) was selected to provide at least 80% power to detect a statistically significant difference (2-fold change from baseline to 12 month) in either dose group for the primary endpoints based on a paired t test with Bonferroni adjustment for multiplicity (α=0.05/6 endpoints/2 doses=0.004, CV=90%), with 10% loss to follow-up rate.

#### Fatty acid profiles

Descriptive statistics were used to summarize patient and disease characteristics. Changes in continuous outcomes (n-3 PUFAs: EPA, DHA, total n-3 PUFAs and n-6 PUFAs: LA, AA, total n-6 PUFAs) from 0-12 months are of primary interest. Mixed effects models for repeated measures (MMRM) with compound symmetry covariance matrix was utilized to model the effect of study arm over time with main effects and the interaction term (time*study arm) as fixed effects with subjects as a random effect. All models for tissue, erythrocytes, and plasma were adjusted for Baseline (T =0) values for age, BMI, WHR, and baseline value of the covariate of interest. All covariates chosen for adjustment were either scientifically significant (e.g., age,^17^ BMI,^5^ baseline value,^5,18^ and WHR^19^), univariately statistically significant, or improved the overall model performance through Akaike Information Criterion (AIC). The Benjamini-Hochberg (BH) procedure^20^ was implemented on the overall interaction F-Test to control the false-discovery rate and limit outcomes of interest amongst tissue, erythrocytes, and plasma with a family-error rate of α =0.05. Specific contrast statements were implemented for specific comparisons of interest in this study (e.g., Difference in change from baseline to 12 months between arms). Pairwise comparisons were performed, using Tukey-Kramer adjustment for multiple comparisons. All calculations were performed using SAS version 9.4 (Statistical Analysis System, RRID:SCR_008567).

#### Lipid mediator data

We conducted MMRM for each outcome similarly as defined above. All models for free and alkaline releasable esterified metabolites were assessed for normality using an Anderson Darling test lending to models based on the log10-transformed values of the outcomes of interest. The potential for storage dependent changes in metabolites were evaluated by testing baseline (T0) samples in a least squares regression model using time from collection to analysis (i.e., storage time) and extraction batch as main effects. As the concentrations of 13-HODE were unaffected by treatment dose or time (p >0.05), measured 13-HODE concentrations were included to control for adipose tissue sampling heterogeneity indicated by replicate analyses. Hence all models were adjusted for baseline (T=0) values for age, BMI, WHR, and baseline value of the covariate of interest, sequencing batch number, time from collection to analysis, as well as their associated log10[13-HODE] to adjust for subsampling heterogeneity. The BH procedure was again used to limit outcomes of interest through controlling the false discovery rate (α =0.05) for the overall interaction effect and contrast statements were implemented as previously described.

#### DNAm data processing and analysis

Samples libraries were generated using a reduced representation bisulfite sequencing (RRBS) protocol and sequenced on an Illumina Novaseq 6000 to an average read depth of 63 million read (minimum depth > 41 million reads; see **Supplementary Methods**). Reads were processed using the nf-core methylseq pipeline version v1.6.1 using the UCSC hg38 reference genome. Trimmed reads were aligned using the Bismark algorithm (version 0.23.0) using the HISAT2 (RRID:SCR_015530) aligner (version 2.2.1). All samples passed QC and incomplete conversion rates were acceptably low (mean=.99%, max=1.55%). DNAm was quantified using Bismark methXtract and averaged for each gene promoter using the hg38 GENCODE refgene gene transcripts, defining the promoter region as 1kb up–– and 1kb down-stream of each transcriptional start site. Only CpGs with at least 10 reads coverage were considered, and DNAm was quantified within each promoter region by taking the mean value of all pass-coverage CpGs. DNAm was summarized for each promoter by taking the average of all samples in each group being compared (i.e., the average of all 12-month, high-dose samples). To control spurious DNAm changes, only promoters with DNAm changes greater than 5% were included in the analysis.

### Data Availability Statement

The human sequence data generated in this study are not publicly available due to patient privacy requirements but are available upon reasonable request from the corresponding author. Other data generated in this study are available within the article and its supplementary data files.

## Results

### Study Cohort

From 2014-2018, women with history of ERPR(−) breast cancer were enrolled at the Ohio State University and City of Hope. A total of 77 women were randomized to ∼5g/d n-3 PUFAs (n =41) or ∼1g/d n-3 PUFAs (n =36) for 12 months (**Fig. 1**). One participant did not receive the allocated intervention (high-dose arm) as a screen failure. There were 59 participants (n =29 ∼5g/d, n =30 ∼1g/d dose) evaluable with at least one follow-up study visit (**Table 1**); 51 participants completed the 12-month intervention (n =26 high-dose, n =25 low-dose). Participant ages ranged from 37 to 75 years. Women self-identified as White (89.8%), Black (8.5%), Asian American (1.7%), and Hispanic (6.8%). Baseline BMI and WHR mean values were elevated at 29.2 kg/m^2^ and 0.83, respectively, without significant difference between study arms (p =0.52 and p =0.43). Eligibility criteria required women to be within 5 years of completion of cancer therapy for ERPR(−)HER2 (+/−) breast cancer, such that the average time from diagnosis to study enrollment was 32 months (6.5 to 66 months).

**Figure 1.**
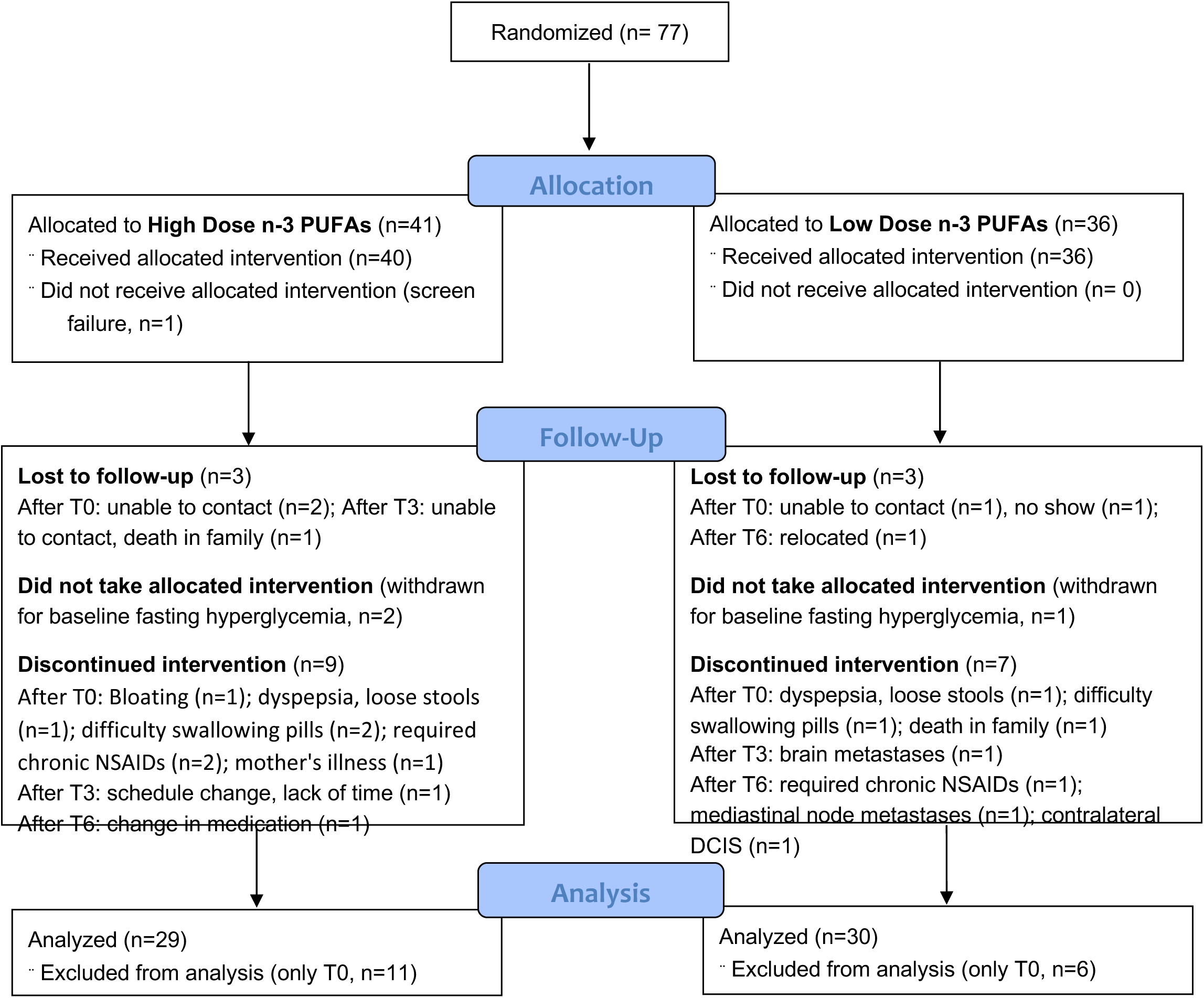
CONSORT Flow diagram.

**Table 1.** Patient characteristics.

| **Table I – Baseline Data<sup>a</sup>** |  | **Total (n=59)** | **High (n=29)** | **Low (n=30)** | **P-Value<sup>b</sup>** |
| --- | --- | --- | --- | --- | --- |
| Age (years) | mean ± SD | 57.1 ± 9.1 | 56.2 ± 9.8 | 57.9 ± 8.5 | 0.50 |
|  | range | 37-75 | 37-75 | 40-75 |  |
| Race, Ethnicity | White | 53 | 25 | 28 | 0.40 |
|  | African American | 5 | 4 | 1 |  |
|  | Asian American | 1 | 0 | 1 |  |
|  | Hispanic | 4 | 2 | 2 |  |
| Weight | mean ± SD | 170.2 ± 33.4 | 173.8 ± 38.9 | 166.8 ± 27.4 | 0.43 |
| BMI | mean ± SD | 29.1 ± 5.2 | 29.3 ± 5.8 | 28.6 ± 4.7 | 0.47 |
| WHR | mean ± SD | 0.83 ± 0.07 | 0.83 ± 0.07 | 0.84 ± 0.06 | 0.37 |
| Tdiagnosis-T0<sup>c</sup> | Months ± SD | 31.8 ± 18.5 | 27.4 ± 15.7 | 34.9 ± 19.9 | 0.11 |
| Stage |  |  |  |  | 0.07 |
|  | 0 | 8 | 5 | 3 |  |
|  | I | 15 | 11 | 4 |  |
|  | II | 31 | 12 | 19 |  |
|  | III | 5 | 1 | 4 |  |
| HER2 overexpression |  |  |  |  | 0.58 |
|  | Negative | 35 | 15 | 20 |  |
|  | Positive | 14 | 8 | 6 |  |
|  | Not tested<sup>d</sup> | 10 | 6 | 4 |  |
| Surgery |  |  |  |  | 0.59 |
|  | Lumpectomy | 39 | 18 | 21 |  |
|  | Mastectomy | 20 | 11 | 9 |  |
| Chemotherapy |  |  |  |  | 0.40 |
|  | None | 11 | 7 | 4 |  |
|  | Adjuvant<sup>e</sup> | 19 | 7 | 12 |  |
|  | Neoadjuvant | 31 | 15 | 16 |  |
| Radiation |  |  |  |  | 0.40 |
|  | None | 12 | 8 | 4 |  |
|  | Lumpectomy | 39 | 18 | 21 |  |
|  | Postmastectomy | 8 | 3 | 5 |  |
| <sup>a</sup> Participants with at least one follow-up study visit.<br><sup>b</sup> Categorical variables tested with Fisher's Exact Test, continuous with 2-sample t-test<br><sup>c</sup> Time from initial diagnosis of ERPR- breast cancer to baseline study visit<br><sup>d</sup> Not tested for DCIS (n=8) and insufficient invasive tumor tissue for testing (n=1)<br><sup>e</sup> 2 participants underwent both neoadjuvant and adjuvant chemotherapy in the low dose group |  |  |  |  |  |

### Adherence, Safety and Tolerability

Adherence was high as assessed by pill counts and patient self-report. Based on pill counts at 3, 6, 9, and 12-month study visits, mean adherence was 91% (n=26), 92% (n=25), 92% (n=25), and 92% (n=21) for the high-dose arm and 94% (n=25), 94% (n=24), 93% (n=24), and 91% (n=21) for the low-dose arm, respectively.

Relative to baseline, ALT, HDL, total cholesterol, and LDL-cholesterol levels did not differ significantly at 0, 6, or 12 months by time or dose (p =NS). Triglyceride levels in the high-dose n-3 PUFA arm decreased from mean values of 111.3 ± 56.89 mg/dL at baseline to 83.1 ± 33.4 mg/dL and 86.6 ± 31.6 mg/dL at 6 months (p =0.01) and 12 months (p =0.008), respectively, without significant change in the low-dose group by Student’s *t* test. Platelet function remained within normal limits in 46 of 51 participants who completed the 12-month intervention. With this test of blood clot formation, elevated closure times (i.e. retarded clot formation rates) were detected in 5 participants at 12 months: 1 in the high-dose arm (184 sec vs ULN of 183 sec) and 4 in the low-dose-arm (ranging from 200-300 sec), with the secondary test indicating an aspirin-like effect. The abnormal PFA-100 results did not correlate with clinically significant bleeding; easy bruising, grade 1 was noted by 10 participants at least once during the study period (4 in the low-dose group, 6 in the high-dose group).

Study treatments were generally well-tolerated. The most frequent adverse events in both arms were gastrointestinal symptoms such as belching, loose stools, flatulence, fishy aftertaste, and nausea, as grade 1 events. Recurrent breast cancer was diagnosed in three participants in the low-dose n-3 PUFA group: 1) mediastinal lymph node metastases; 2) brain metastases; and 3) contralateral breast ductal carcinoma in situ. These adverse events appeared related to disease progression rather than study treatment.

### Fatty acid profiles

Fatty acid profiles were analyzed in breast adipose tissue, erythrocyte membranes, and plasma specimens obtained at 0, 3, 6, 9, and 12 months of high-vs low-dose EPA+DHA. Adipose tissue fatty acid profiles indicate dietary fatty acid intake over years, while erythrocyte and plasma fatty acids reflect fat consumption over the past one to two months and one to two weeks, respectively.^21^ For the primary analysis, n-3 PUFAs EPA, DHA, and total n-3 PUFAs and n-6 PUFAs linoleic acid (LA), arachidonic acid (AA), and total n-6 PUFAs were compared between and within the high-dose and low-dose n-3 PUFA study arms at 0, 3, 6, 9, and 12 months, controlling for variables of age, baseline metabolite values, BMI, and WHR.

In breast adipose tissue, baseline breast adipose fatty acids did not differ significantly between the two groups (**Table 2**). The BH-adjusted interaction effect was significant (p <0.05) for mean percentage of total fatty acids, EPA, DHA, and total n-3 PUFAs. Based on contrast statements, mean percentage of total fatty acids, EPA, DHA, and total n-3 PUFAs differed significantly between the two study arms at 3, 6, 9, and 12 months of treatment, with higher mean EPA, DHA, and total n-3 PUFAs in the high-dose vs low-dose group at each time interval (**Fig. 2A**). LA and AA and total n-6 PUFAs did not demonstrate significant dose*time interaction (p >0.1) in breast adipose tissue, and similarly, no significant difference within or between arms. In addition to significant increases in n-3 PUFAs, we also observed variability between individuals in the overall fatty acid profiles, including n-3 PUFA concentrations, in both study arms (**Fig. 2B**).

**Figure 2.**
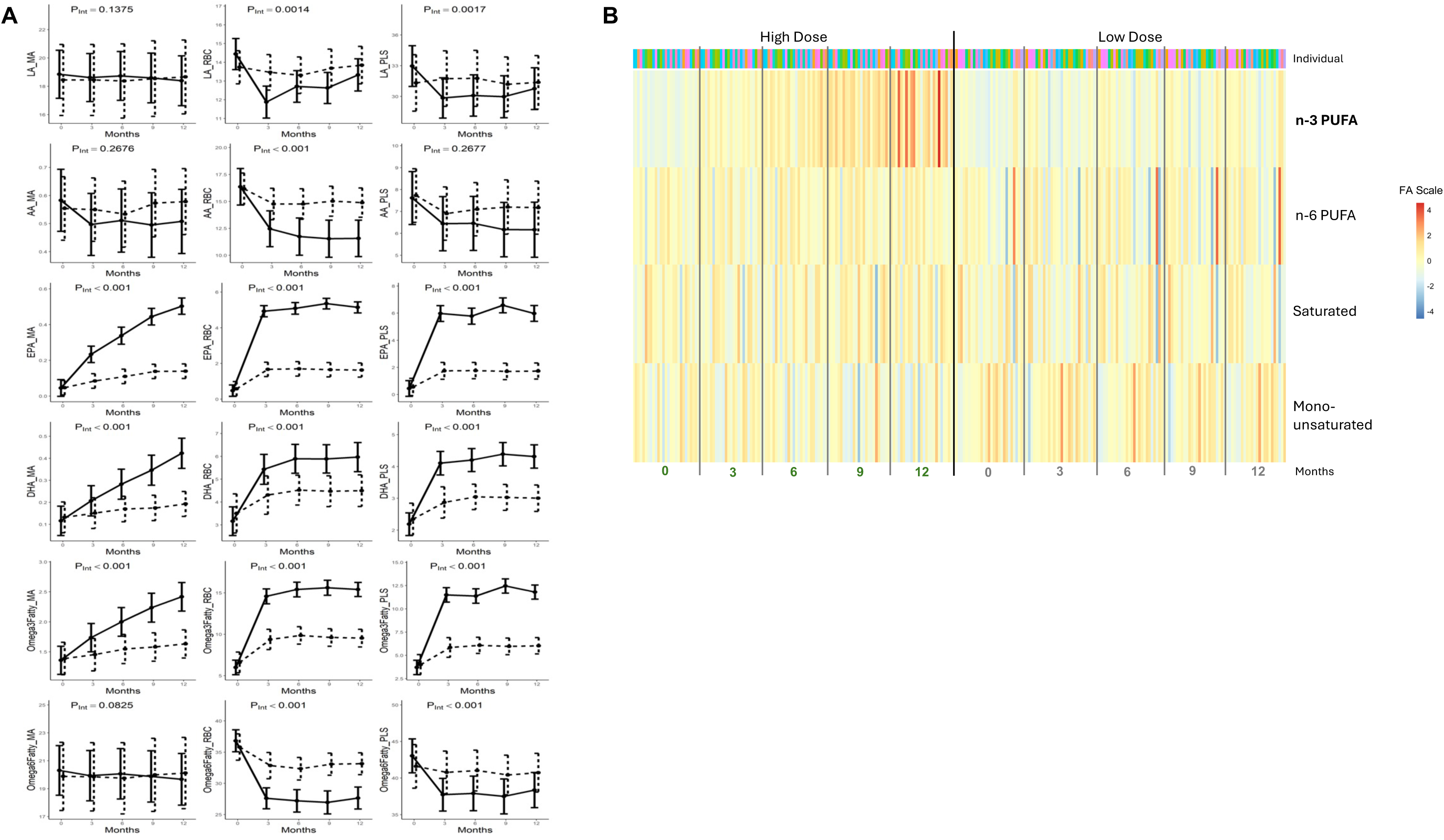
Fatty acid profiles. A) n-3 PUFAs and n-6 PUFAs as mean percentages of total fatty acids in breast adipose (MA), erythrocyte membranes (RBC), and plasma (PLS) at 0, 3, 6, 9, and 12 months of 5g/d and 1g/d, solid and dashed lines respectively. Mean estimates for linoleic acid (LA), arachidonic acid (AA), eicosapentaenoic acid (EPA), docosahexaenoic acid (DHA), total n-3 PUFAs, total n-6 PUFAs with SD error bars. B) Fatty acid heat map. Breast adipose total n-3 PUFAs, n-6 PUFAs, SFAs, and MUFAs sorted by samples (n=51) grouped by time (0, 3, 6, 9, 12 months) within each arm. Fatty acid values were mean centered within each fatty acid and across both arms to show how each fatty acid changed during treatment. Note: fatty acid groups are not depicted proportional to scale/percentage.

**Table 2.** Breast adipose fatty acids 0-12 months high and low dose n-3 PUFAs.

| FattyAcid<sup>1</sup> | Arm<sup>2,3</sup> | 0 months | 3 months | 6 months | 9 months | 12 months |
| --- | --- | --- | --- | --- | --- | --- |
| 12:0 | high | 0.48 (.26) | 0.43 (.15) | 0.45 (.17) | 0.47 (.15) | 0.46 (.17) |
|  | low | 0.52 (.29) | 0.50 (.24) | 0.52 (.23) | 0.54 (.28) | 0.56 (.29) |
| 14:0 | high | 2.56 (.44) | 2.57 (.45) | 2.58 (.40) | 2.6 (.40) | 2.59 (.44) |
|  | low | 2.59 (.72) | 2.52 (.68) | 2.67 (0.77) | 2.57 (.70) | 2.59 (.68) |
| 16:0 | high | 21.25 (1.51) | 21.12 (1.50) | 21.18 (1.48) | 21.39 (1.58) | 21.46 (1.50) |
|  | low | 21.29 (1.62) | 21.05 (1.60) | 21.36 (1.65) | 21.06 (1.67) | 21.01 (1.58) |
| 16:1n7 | high | 3.55 (1.00) | 3.66 (1.10) | 3.44 (1.14) | 3.44 (1.08) | 3.47 (1.16) |
|  | low | 3.5 (1.08) | 3.67 (1.12) | 3.66 (1.23) | 3.62 (1.19) | 3.69 (1.16) |
| 18:0 | high | 4.26 (.94) | 4.17 (1.00) | 4.40 (.97) | 4.39 (.93) | 4.43 (0.96) |
|  | low | 4.04 (.87) | 4.07 (1.06) | 4.06 (.87) | 4.06 (.97) | 3.86 (.85) |
| 18:1n-9 | high | 43.43 (2.01) | 43.49 (1.91) | 43.06 (1.63) | 42.89 (2.09) | 42.79 (2.28) |
|  | low | 43.81 (2.15) | 44.01 (2.31) | 43.56 (2.06) | 43.60 (2.07) | 43.7 (2.16) |
| 18:1n-7 | high | 2.19 (.30) | 2.20 (.26) | 2.18 (.29) | 2.10 (.24) | 2.12 (.27) |
|  | low | 2.24 (.35) | 2.26 (.34) | 2.25 (0.31) | 2.26 (.36) | 2.19 (.32) |
| 18:2n-6 | high | 18.82 (1.71) | 18.57 (1.93) | 18.70 (1.94) | 18.55 (1.85) | 18.34 (1.67) |
|  | low | 18.48 (2.52) | 18.39 (2.52) | 18.35 (3.15) | 18.61 (2.98) | 18.72 (2.96) |
| 18:3n-6 | high | 0.07 (.02) | 0.07 (.02) | 0.07 (.02) | 0.07 (.02) | 0.07 (.02) |
|  | low | 0.07 (.02) | 0.07 (.02) | 0.07 (.02) | 0.07 (0.01) | 0.07 (.01) |
| 18:3n-3 | high | 0.98 (.16) | 1.0 (.15) | 1.01 (.16) | 1.02 (.13) | 1.01 (.14) |
|  | low | 0.99 (.21) | 0.98 (.19) | 1.00 (.20) | 0.99 (.25) | 1.01 (.25) |
| 18:4n-3* | high | 0.01 (.01) | 0.02 (.02) | 0.02 (.02) | 0.02 (.02) | 0.03 (.02) |
|  | low | 0.01 (.01) | 0.01 (.01) | 0.01 (.01) | 0.01 (.01) | 0.01 (.01) |
| 20:0 | high | 0.12 (.05) | 0.11 (.06) | 0.12 (.06) | 0.12 (.05) | 0.12 (.05) |
|  | low | 0.11 (.04) | 0.11 (.05) | 0.01 (.04) | 0.11 (.05) | 0.11 (.04) |
| 20:1Δ11n9 | high | 0.52 (.11) | 0.53 (.06) | 0.54 (.06) | 0.50 (.12) | 0.51 (.07) |
|  | low | 0.53 (.09) | 0.54 (.11) | 0.52 (.10) | 0.53 (.09) | 0.50 (.12) |
| 20:2n-6 | high | 0.25 (.05) | 0.24 (.05) | 0.25 (.04) | 0.24 (.04) | 0.23 (.04) |
|  | low | 0.25 (.05) | 0.25 (.06) | 0.23 (.06) | 0.24 (.06) | 0.25 (.06) |
| 20:3n-6 | high^ | 0.30 (.10) | 0.28 (.09) | 0.28 (.07) | 0.26 (.09) | 0.27 (.08) |
|  | low | 0.30 (.10) | 0.29 (.11) | 0.29 (.09) | 0.31 (.11) | 0.30 (.10) |
| 20:4n-3* | high^ | 0.03 (.01) | 0.05 (.01) | 0.06 (.02) | 0.07 (.02) | 0.08 (.03) |
|  | low^ | 0.03 (.01) | 0.03 (.01) | 0.04 (.01) | 0.04 (.02) | 0.04 (.01) |
| 20:4n-6 | high | 0.59 (.20) | 0.50 (.13) | 0.51 (.14) | 0.51 (.14) | 0.51 (.16) |
|  | low | 0.55 (.17) | 0.54 (.20) | 0.54 (0.14) | 0.57 (.21) | 0.57 (.21) |
| 20:5n-3* | high^ | 0.05 (.04) | 0.24 (.09) | 0.34 (.15) | 0.46 (.17) | 0.51 (.30) |
|  | low^ | 0.05 (.03) | 0.08 (.05) | 0.11 (.05) | 0.14 (.08) | 0.14 (.06) |
| 22:4n-6 | high | 0.21 (.07) | 0.19 (.08) | 0.19 (.07) | 0.19 (.07) | 0.17 (.07) |
|  | low | 0.23 (.01) | 0.22 (.11) | 0.21 (.08) | 0.22 (.10) | 0.23 (.09) |
| 22:5n-3* | high^ | 0.17 (.05) | 0.24 (.06) | 0.29 (.07) | 0.34 (.08) | 0.38 (.12) |
|  | low^ | 0.19 (.06) | 0.21 (.07) | 0.23 (.06) | 0.24 (.07) | 0.25 (.08) |
| 22:5n-6 | high | 0.03 (.01) | 0.03 (.01) | 0.03 (.02) | 0.03 (.01) | 0.03 (.01) |
|  | low | 0.03 (.01) | 0.03 (.03) | 0.03 (02) | 0.03 (.01) | 0.03 (.02) |
| 22:6n-3* | high^ | 0.12 (.08) | 0.21 (.06) | 0.29 (.09) | 0.35 (.11) | 0.43 (.23) |
|  | low | 0.13 (.08) | 0.15 (.07) | 0.18 (.08) | 0.17 (.07) | 0.19 (.09) |
| Total n-3* | high^ | 1.36 (.24) | 1.76 (.26) | 2.01 (.34) | 2.26 (.37) | 2.43 (.70) |
|  | low^ | 1.40 (.28) | 1.47 (.31) | 1.57 (.35) | 1.59 (.37) | 1.64 (.38) |
| Total n-6 | high^ | 20.28 (1.80) | 19.88 (2.02) | 20.03 (1.98) | 19.85 (1.91) | 19.62 (1.69) |
|  | low | 19.90 (2.48) | 19.79 (2.54) | 19.73 (3.19) | 20.05 (3.03) | 20.16 (3.00) |
| Total MUFA | high | 49.69 (2.38) | 49.87 (2.23) | 49.22 (2.16) | 48.93 (2.61) | 48.89 (2.69) |
|  | low | 50.15 (2.47) | 50.48 (2.54) | 49.99 (2.40) | 50.01 (2.49) | 50.08 (2.57) |
| Total SFA | high | 28.68 (2.17) | 28.49 (2.44) | 28.74 (2.20) | 28.96 (2.44) | 29.06 (2.28) |
|  | low | 28.54 (2.56) | 28.26 (2.59) | 28.71 (2.65) | 28.35 (2.46) | 28.12 (2.44) |
| n-6:n-3* | high^ | 15.29 (2.66) | 11.54 (1.97) | 10.31 (2.42) | 9.07 (2.14) | 8.79 (2.88) |
|  | low^ | 14.55 (2.44) | 13.91 (2.92) | 12.94 (.48) | 13.12 (2.94) | 12.75 (2.43) |

<sup>1</sup>Fatty acids as percentage of total fatty acids (weight %) with standard deviation (SD)<sup>2</sup>number of high dose subjects at 0, 3, 6, 9, and 12 months: 27, 25, 26, 25, 25<sup>3</sup>number of low dose subjects at 0, 3, 6, 9, and 12 months: 28, 29, 27, 24, 25
\*p-values<.001, 0-12 months between arms except for 18:4n-3 p=.009
^p-values<.001, 0-12 months within arm. 22:6n-3 low dose arm p=.0016

BH-adjusted erythrocyte membrane EPA, DHA, total n-3 FAs, total n-6 FAs, LA, and AA differed significantly following ∼5g/d versus ∼1g/d n-3 PUFA supplementation (**Fig. 3A,B**, **Supplementary Table S2**), with significantly higher mean percentage EPA, DHA, and total n-3 PUFAs in women in the higher dose group at 3, 6, 9, and 12 months that plateaued at 3 to 6 months. The omega-3 index (sum of EPA and DHA as weight percent of total FAs in erythrocytes) increased from a baseline of 3.63% and 4% in high– and low-dose groups, respectively, to peak of ∼ 11% and 6%, respectively, at 6 months without further significant increase at 9 or 12 months (**Fig. 3C, Table S2**). LA, AA, and total n-6 PUFAs in erythrocytes decreased significantly by treatment arm, with lower mean LA and AA as percentage of total fatty acids in the high-dose relative to low-dose arm (**Fig. 2A**). Mean percentage of LA significantly decreased for the high-dose but not low-dose arm; mean AA % decreased significantly in both arms but with greater reduction in women on high-dose supplementation.

**Figure 3.**
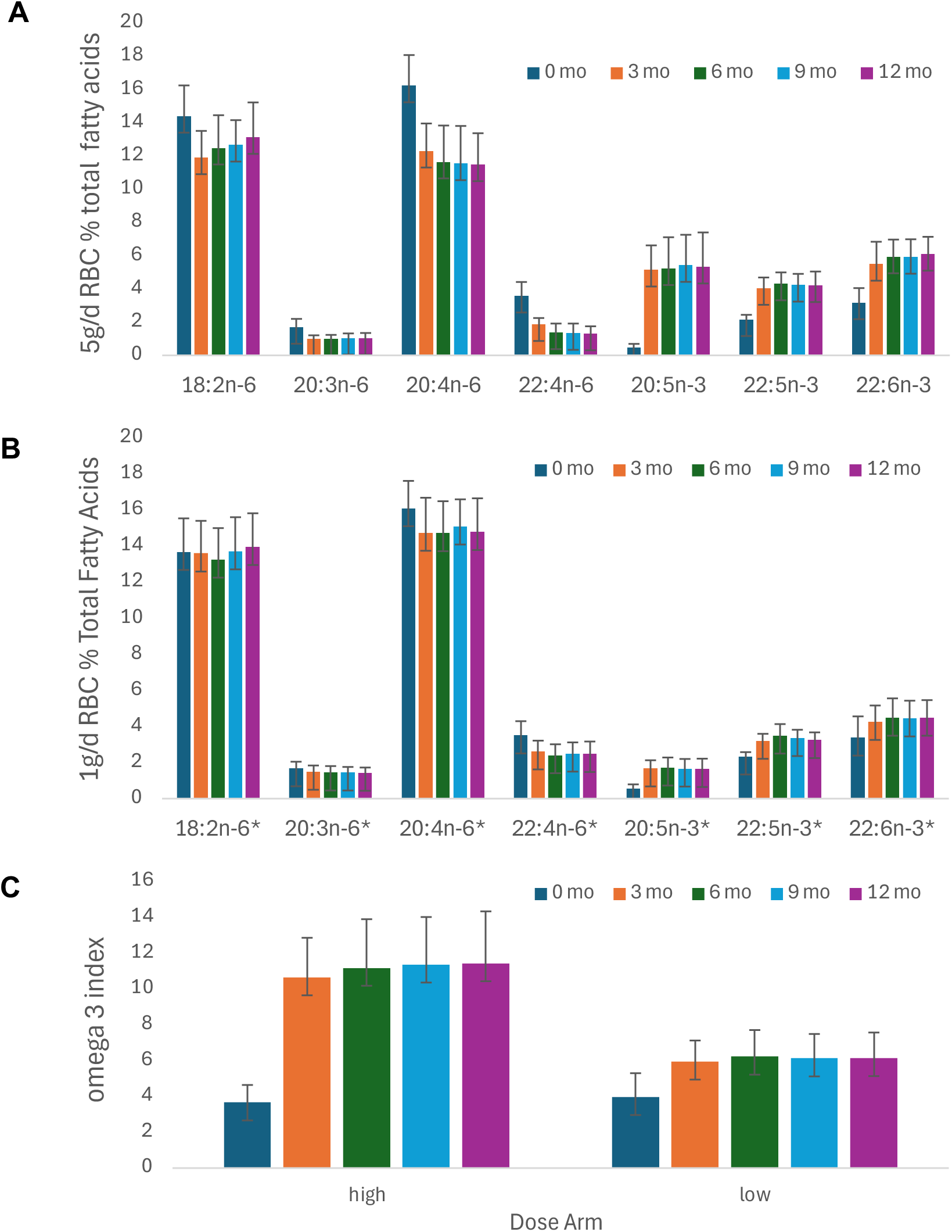
Erythrocyte membrane PUFAs, Fatty acids expressed as percentage of total fatty acids (weight %) with standard deviation (SD) for n-6 PUFAs 18:2n-6 (LA), 20:3n-6 (?), 20:4n-6 (AA), 22:4n-6 (?) and n-3 PUFAs 20:5n-3 (EPA), 22:5n-3 (DPA), and 22:5n-3 (DHA) at 0, 3, 6, 9, and 12 months for. **A)** 5g/d, n=32 and B) 1g/dn=?. C) omega-3 index as sum of EPA+DHA content of erythrocytes as percentage total fatty acids (weight %). *p-values<.001, 0-12 months between arms except 18:2n-6=.032 by MMRM.

As in erythrocytes, plasma EPA, DHA, and total n-3 PUFAs increased whereas LA and total n-6 PUFAs decreased significantly as relative percentage of total fatty acids in BH-adjusted models with both treatment arms, stabilizing at 3 to 6 months (**Fig. 2A**). The changes in relative percentage were significantly higher in the high-dose vs low-dose groups 3, 6, 9, and 12 months.

### Breast adipose fatty acid metabolite analyses

Breast adipose tissue samples obtained at 0, 6, and 12 months were analyzed in a subset of randomly selected study participants (n =28) for non-esterified and alkaline-releasable esterified oxylipin levels. Samples were processed and analyzed for non-esterified oxylipins in breast adipose tissue at all time points; however, due to limited sample avalability, the final analysis cohort for the esterified oxylipin data set included only 22/28 complete sets of samples for all time points and remaining samples matched for two (n =5) and 1 (n =1) time points (**Supplementary Fig. S1**).

Initial analysis yielded 41 esterified oxylipins with significant dose*time interactions (p <0.05), and the BH adjustment reduced that to 34 significant outcomes that we identified to investigate further. Of those, they were primarily EPA-(n =15) and DHA-derived (n=13), with fewer AA-(n =1), LA-(n =3) and ALA-derived (n=2) oxylipin changes (**Fig. 4A, Supplementary Table S3**). The changes in oxylipin level correlated positively with dose for n-3 PUFA metabolites and negatively for n-6 PUFA metabolites.

**Figure 4.**
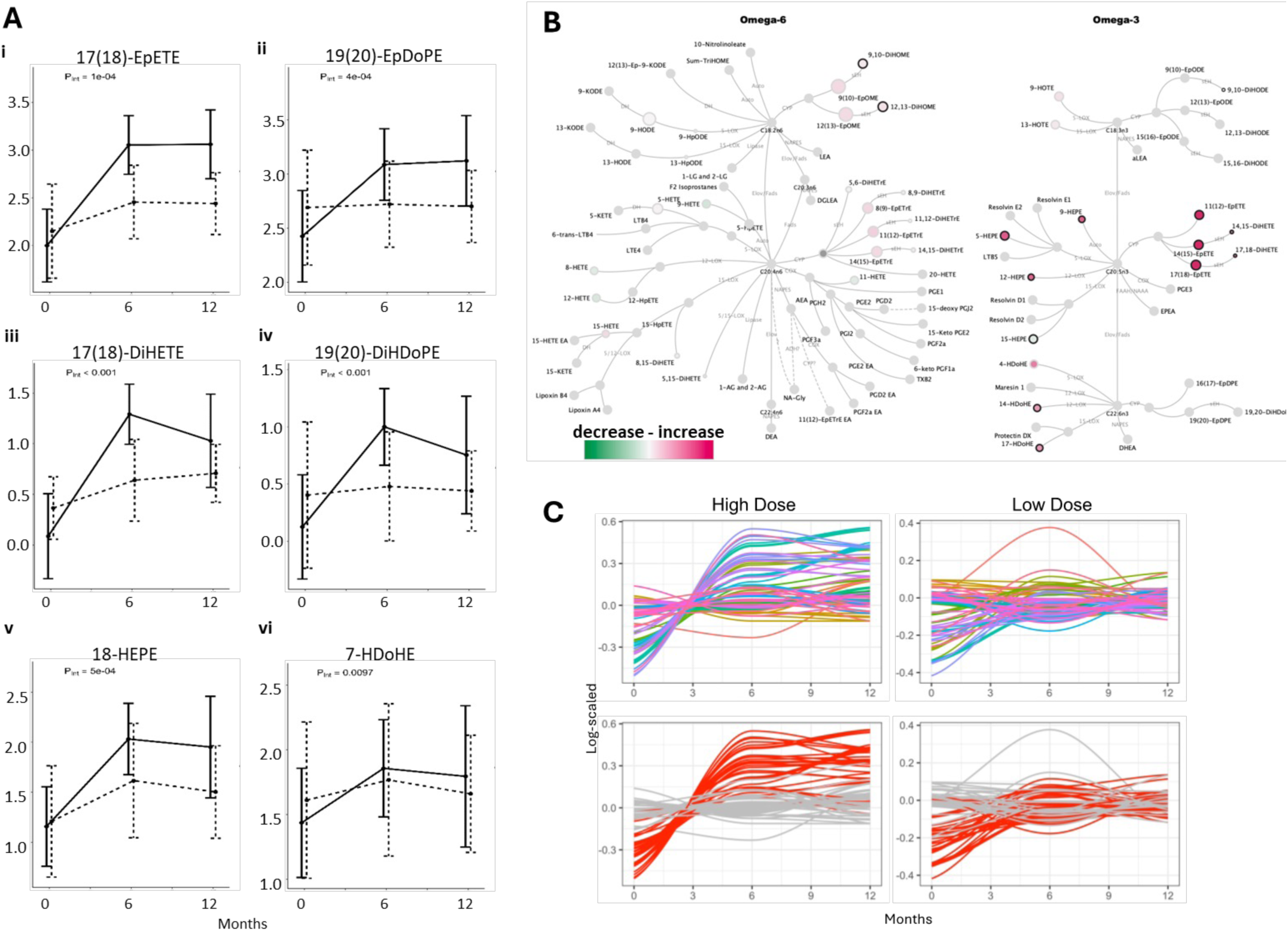
Oxylipins in breast adipose esterified pools. A) Representative oxylipin epoxides (Panel a and b), diols (Panels c and c), and alcohols (Panels e and f). Results are adjusted means of metabolite concentrations by n-3 PUFA dose and treatment time interactions MMRM and BH correction. P-values of interactions are shown along with the r2 of the complete model. B) Metabolite changes in PUFA pathways. The color scale and the point size (larger node has larger change) show the difference in the high dose 12-vs 0-month timepoints for esterified oxylipins. A black border indicates a significant difference was observed over n-3 PUFA treatment. Solid light grey nodes indicate features not tested. C) Summary of all oxylipin metabolites shows individual participants by study arm over time (multicolor lines). Bottom panel shows significant differences in red and nonsignificant in grey.

Analysis of esterified oxylipin profiles in breast adipose tissue samples for dose by time effects demonstrated significant increases primarily in DHA– and EPA-derived oxylipins between 0 to 12 months, with significantly higher levels in the high-dose relative to low-dose treatment arm over time (**Fig. 4B**). By covariate-adjusted analysis, both treatments yielded increased mean concentrations of EPA and DHA epoxides, diols, and alcohols (**Table S3**), which correlated with the parent fatty acids EPA and DHA in breast adipose tissue. In addition to the statistically significant increases in specific EPA– and DHA-derived oxylipins, concentrations of several ALA-, LA-, and AA-derived oxylipins decreased significantly with n-3 PUFA treatment from 0-12 and/or 6-12 months (**Table S3**).

The non-esterified pool of n-3 PUFA derived alcohols in breast adipose tissue also increased significantly from 0 to 12 months, with dose-dependent effects for two oxylipins: 9-HEPE and 18-HEPE. With the BH procedure, non-esterified 9-HEPE and 18-HEPE demonstrated significant interaction effects, with increased concentration from 0 to 12 months. Non-esterified epoxides and diols were not detected at significantly different levels by treatment or time.

In general, the fatty acid metabolites affected by EPA+DHA supplementation increased from baseline to 6 months without further significant increase from 6 to 12 months, with the higher dose showing more consistent and substantial effects. Metabolite changes plotted for each participant demonstrate individual variability (**Fig. 4C**).

### DNAm in breast adipose tissue

The canonical role of DNAm in the CG-rich promoter regions is to regulate the expression of associated genes: methylated CG-rich promoter regions are generally associated with gene silencing and unmethylated promoter regions with gene expression. We investigated DNAm in breast adipose samples obtained 0, 6, and 12 months (n =7 high-dose, n =10 low-dose) using RRBS, a method that primarily interrogates CG-rich promoter regions. For this subset of samples, changes in fatty acids were consistent with the full study analyses.

Our prior study of DNAm in PBMCs of high-risk women treated with high-dose ∼5 g/day EPA+DHA showed marked enrichment of hypermethylation of inflammation-related pathways at 6 months by RRBS analyses.^22^ To similarly investigate DNAm in breast adipose tissue for inflammation effects, we first assessed whether DNAm changes were observed in inflammation-related pathways (n =10, Wikipathway database,^23^ **Supplementary Table S4.1**). For each pathway, DNAm of each gene promoter region was quantified for each sample; mean promoter DNAm was then compared between high-dose versus low-dose samples obtained at 12 months of n-3 PUFA treatment. For promoters with an effect size >5% DNAm change, the number of hyper-to hypo-methylated gene promoters in each pathway were compared to determine the n-3 PUFA treatment effect. Although DNAm changes were observed in inflammation-related genes (**Table S4.2**), none of the inflammation-related pathways were enriched for concerted DNAm changes at 12 months of n-3 PUFAs.

Applying the same approach, we additionally assessed whether DNAm changes occurred in fatty acid-or metabolism-related pathways^23^ (n =22, **Fig. 5**, **Table S4.3**). Using a binomial test to determine whether the pathway was enriched for hyper-or hypo-methylation, two pathways were significantly hypermethylated after n-3 PUFA treatment (**Fig. 5, Supplementary Fig. S2**): 1) sterol regulatory element-binding proteins (SREBP) signaling pathway which involves the transcriptional regulation of lipid homeostasis including cholesterol, triglyceride, and fatty acid synthesis^24^ and has hypermethylated promoters located primarily in genes of the endoplasmic reticulum (ER) as well as AMPK and LPIN1 genes; 2) orexin receptor pathway which involves key metabolic and physiologic processes such as triglyceride production and has hypermethylation across the pathway in both diverse metabolism-related genes (e.g., LEP, COX7A1, ALPP, TH) and inflammation-related gene MAPK14.

**Figure 5.**
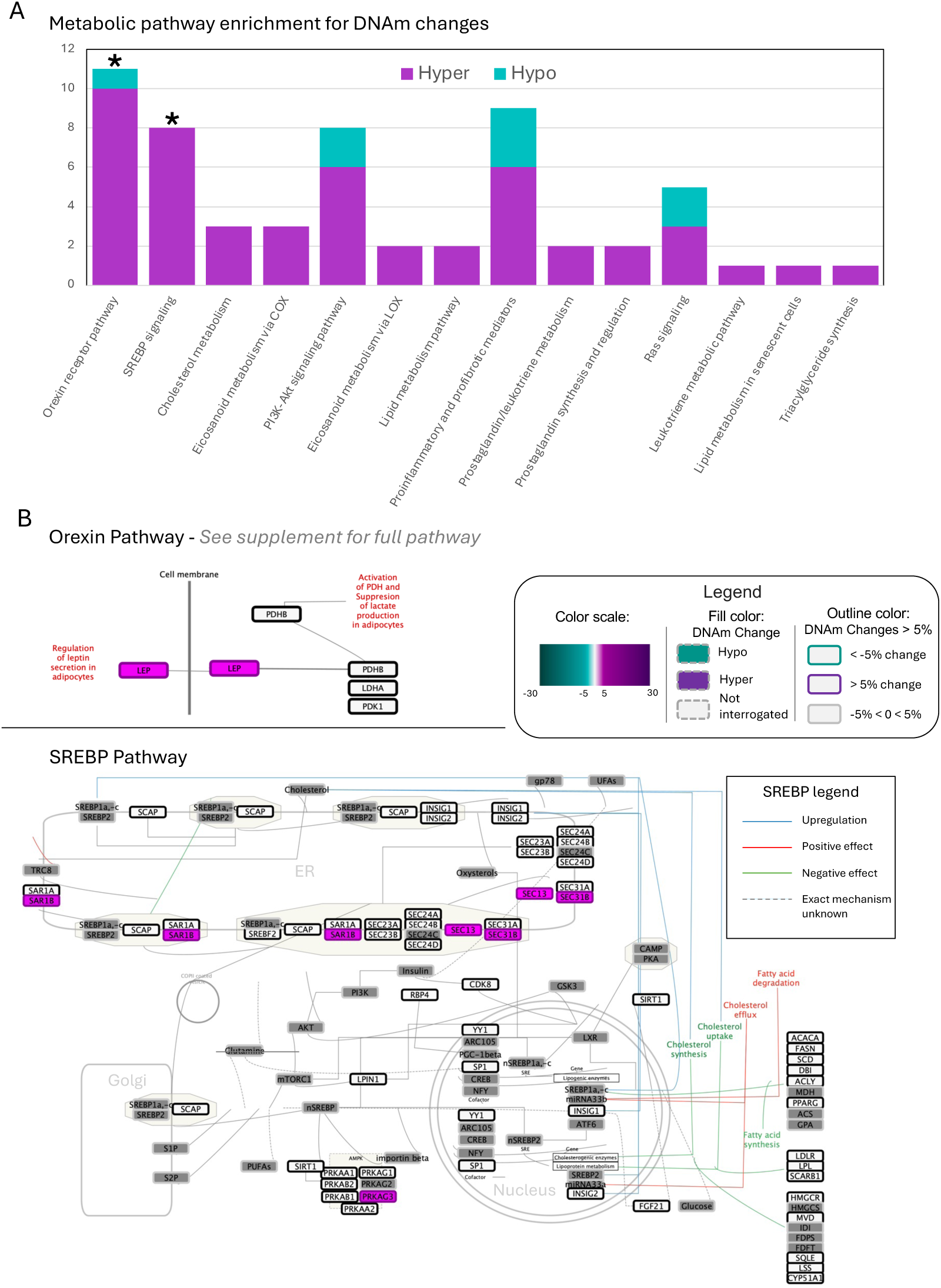
DNAm analysis of metabolism pathways. A) DNAm of the gene promoters in the metabolism-related pathways were compared between the high dose vs the low dose arms at the final 12-month timepoint. The number of hypermethylated (increased DNAm in the high dose arm) vs hypomethylated promoters were tested for enrichment using a binomial test (FDR p-value < 0.05). B) Two pathways were determined to be significantly hypermethylated. DNAm was visualized in each significant pathway to explore how the observed DNAm changes might affect the function of the pathway. See **Fig. S2** for additional detail of the orexin pathway.

## Discussion

With this randomized, double-blind dose-response trial of high-dose n-3 PUFAs in women with ERPR(−) breast cancer, we demonstrate dose and time effects of EPA+DHA supplementation on breast adipose fatty acids and associated metabolites. The study intervention was well-tolerated without attributable serious adverse events. Additionally, withdrawals did not generally relate to side effects of either EPA+DHA dose. Consistent with prior reports, triglyceride levels decreased in women in the high-dose but not low-dose arm; women in the low-dose arm reached a maximum omega 3 fatty acid index of ∼ 6% whereas those in the high-dose arm reached a stable level of ∼11% at 6 to 12 months. Elevated clotting time (PFA-100) at 12 months (n =5) did not appear related to study dose, with secondary test indicative of aspirin-like effects possibly related to intermittent NSAID use.

For women treated with ∼5g/d, the mean percentage of n-3 PUFAs continued to increase in breast adipose tissue without apparent plateau at 12 months. Individual variability in EPA and DHA incorporation into breast adipose tissue over 12 months is evident; for example, 6 of 25 participants in the high-dose group showed increases at 9 to 12 months (data not shown). In contrast, with 1g/d, breast adipose DHA and EPA content by mean values stabilized at 6 to 12 months at levels significantly below the high-dose arm by over 2-to 3-fold, respectively.

For most EPA– and DHA-derived metabolites, concentrations stabilized at 6 to 12 months in both study arms, reaching a significantly higher plateau level in the high-dose vs low-dose group. Despite continuing increases in breast adipose EPA and DHA as mean percentage of total fatty acids in the high-dose group, most of the EPA-and DHA-derived metabolites did not concordantly increase in adipose tissue. These results suggest saturation kinetics for EPA– and DHA-derived oxylipins in adipose tissue, possibly due to metabolizing enzyme pathway capacity and/or competition related to relative n-3 PUFA and n-6 PUFA content. In breast adipose tissue, unlike erythrocyte and plasma specimens, increased n-3 PUFA content by mean percentage total fatty acids (**Table 2**) was not associated with concordant reduction in n-6 PUFAs.

Ostermann et al. reported linear increases in EPA– and DHA-derived plasma oxylipins that loosely tracked parent fatty acid changes at 3 and 12 months for intakes up to ∼13g EPA+DHA/week.^6^ Moreover, oxylipin increases were more pronounced in the first 3 months with an apparent leveling of the response towards 12 months.^6^ Our breast adipose tissue metabolite dose response suggests that similar yet unique behaviors are at work in this tissue and raises the possibility that intermittent n-3 PUFA dosing schedules and/or complementary dietary intake might prove sufficient to maintain a specified level of oxylipin(s) once achieved.

The oxylipin profile in breast adipose tissue is notable for greater number of esterified vs unesterified EPA– and DHA-derived metabolites following n-3 PUFA supplementation, consistent with prior reports of oxylipin analyses of plasma samples of participants treated with 12 weeks of long chain n-3 PUFAs.^18,25^ The esterified forms may serve an active or passive reservoir of these compounds. For instance, if oxylipin release is actively and specifically engaged during a biochemical response (e.g. PLA2-dependent release from phospholipid pools), this reservoir may be an integral part of the signaling capacity of a cell. On the other hand, if released during the normal harvesting of lipids within a given pool (e.g. fatty acid release from triglycerides to support energy demands), the pool level may provide regulation of the tonal balance of signaling pathways in which oxylipins participate. Unesterified epoxides and diols were not detected at significantly different levels by dose or time, suggesting that the intensity of the active signaling pool is not altered by dietary intake and tonal regulation clears the non-esterified forms from breast adipose when released. In initial analyses without covariate analyses aside from 13-HODE normalization, esterified HETEs decreased significantly following n-3 PUFA treatment, with greater reduction in the high-dose vs low-dose groups; however, the reductions were not significant following BH correction to FDR 5% (**Table S3**) or 20% (data not shown). Although breast adipose AA-derived HETEs did not decrease significantly, n-3 PUFA-derived LOX oxylipins increased significantly with potential dilutional effect on the HETEs and competition for downstream effects. Given the significantly decreased n-6 PUFAs in plasma but not breast adipose tissue following n-3 PUFA treatment as percentage of total fatty acids (**Fig. 2**), plasma oxylipin profiling might also reveal changes in n-6 PUFA-derived oxylipin species as reported by others.^6,26^

The increase in esterified EPA-derived metabolites, such as 18-HEPE, and DHA-derived metabolites, such as 14-HDoHE, may provide useful measures of n-3 PUFA-mediated anti-inflammatory potential in target organs such the breast where adipose tissue is a key component of the tumor microenvironment. Circulating and tissue-based oxylipins derived from EPA, DHA may play a role in amelioration of lipid-mediated anti-inflammatory signaling in other solid tumors such as suggested by preclinical studies of lung carcinoma, melanoma, and colon cancer.^27–29^

We also explored the potential association of DNAm changes with n-3 PUFA metabolism, identifying DNA promoter hypermethylation in the SREBP and orexin signaling pathways. In general, upregulation of both identified pathways has been implicated in cancer growth and development. SREBP signal transduction is fundamentally involved in fatty acid, cholesterol, and triglyceride metabolism. In rodent models, dietary PUFAs suppress the proteolytic activation of hepatic SREBP-1c and associated autoregulatory feedback loop, a critical element in the regulation of triglyceride synthesis.^30,31^ Our findings are consistent with an SREBP-1 dependent mechanism for triglyceride reduction via high-dose n-3 PUFAs. Regarding breast cancer, elevated SREBP1 expression has been associated with poor prognosis and enhanced cancer growth.^32^ Notably, dyslipidemia has been linked to poorer outcomes for women with TNBC,^33,34^ for which n-3 PUFA supplementation is recommended.^35^ Downregulation of the SREBP pathway, which was hypermethylated by ∼5g/d n-3 PUFA treatment, has been linked to cancer cell growth inhibition and identified as a potential therapeutic target in cancer in general.^36–39^ The orexin pathway acts through activation of the mammalian target of rapamycin (mTOR) and Akt (i.e., protein kinase B) signaling cascades.^40^ Given the implication of the PI3K/Akt/mTOR pathway in nearly all aspects of breast cancer growth and development,^41^ the ability of orexin signaling to activate these cascades may provide a link between diet and cancer.^40^ In addition, hypermethylation of LEP as noted in the orexin pathway suggests potential reduction in leptin signal transduction with downstream effects that inhibit the development and progression of breast cancer.^42,43^ Taken together, EPA+DHA supplementation, at a dose sufficient to downregulate adverse metabolic signaling conducive to ERPR(−) breast cancer development and progression, may represent an eminently feasible and tolerable preventive treatment for this high-risk breast cancer sub-type.

Limitations of the study include the necessity for correlative oxylipin and DNAm analyses in a subset of participants to accommodate constraints of time and resources resulting from laboratory closures and personnel changes during the COVID19 pandemic. Time elapsed between collection and analysis of biospecimens may have also yielded altered oxylipin profiles despite storage at –80°C. Further, breast adipose tissue is comprised of different cell types with potential for variability between and within individuals. Challenges of high risk of recurrence and cancer treatment-related comorbidities, as well as burden of study requirements, affected eligibility and accrual; further, study attrition was higher than anticipated although without specific pattern (**Fig. 1**). Lastly, although our study population reflects the racial and ethnic diversity of the two participating sites, a larger multi-site trial is warranted to determine benefit in different racial and ethnic groups; clinical trials to date indicate potential for greater benefit in certain subgroups, as for African Americans in the Multi-Ethnic Study of Athersclerosis (MESA) with inverse association of higher EPA, DHA with lower cardiovascular disease risk,^44^ which may prove relevant for ERPR(−)HER2(−) breast cancer which disproportionately affects African American women vs other racial/ethnic groups.^45^

This clinical trial is the first to link a dietary intervention with metabolic and epigenetic changes in women with ERPR(−) HER2(+/−) breast cancer, which are high-risk molecular subtypes for which preventive and long-term adjuvant strategies are lacking. High-dose EPA+DHA supplementation at ∼5g/d may offer protective benefits via increased breast adipose EPA-DHA-derived oxylipin levels and DNA hypermethylation of pro-carcinogenic signaling pathways. These results will inform the design of future breast cancer prevention trials of dietary n-3 PUFAs.

## Acknowledgements

We gratefully acknowledge the time, effort, and commitment of the study participants. We thank all the clinic staff, clinicians, and clinical trials team involved in this study at City of Hope National Medical Center/COHCCC and Stefanie Spielman Comprehensive Breast Center/OSUCCC. We acknowledge Dr. Joanne L. Lester and Ms. Maria V. Johnson for invaluable assistance with the conduct of the clinical trial and data management. This clinical trial was funded by R01 CA164019 (LDY). Additional support was provided by USDA Project 2032-51530-025-00D (JWN). The USDA is an equal opportunity employer and provider.

