## Supplemental files for "Randomized dose-response trial of n-3 fatty acids in hormone receptor negative breast cancer survivors—impact on breast adipose oxylipin and DNA methylation patterns"

**Supplementary Methods:**

***DNAm library generation and sequencing***

DNA was extracted from breast FNA samples using Gentra Puregene DNA extraction kit (Qiagen); samples were required to have at least 50ng of material. We modified the previously published protocol^1^ and devolved a streamlined RRBS sequencing library preparation method as follows. Genomic DNA was digested with MspI at 37°C for overnight. In the same reaction tube, ends of the DNA fragments were filled in and a 3′ adenosine was added with Klenow Fragment (3′→5′ exo-minus), then followed by the ligation to add the methylated universal adaptors (IDT xGen™ Stuby Adapter, Cat10005924). The ligation products were purified with 2.0x Ampure XP beads (Beckman Coulter, Cat A63881), followed by bisulfite conversion with EZ DNA Methylation kit (Zymo Research, Cat D5001) and the final library was amplified with Pfu Turbo Cx Hotstart DNA Polymerase (Agilent, Cat600414) by using IDT UDI primers (IDT xGen™ UDI Primer Paris, Cat 10005922). The final libraries were purified with 1.2X Ampure XP beads. The library QC was done with Agilent Tapestation D5000 tape and quantified by Qpcr. RRBS-seq libraries were sequenced on Illumina on the NovaSeq 6000 platform with S4 reagent kit v1.5 (300 cycles). Real-Time Analysis (RTA) v3.4.4 software was used for base calling and Illumina bcl2fastq (RRID:SCR_015058) to convert base call (BCL) files into FASTQ files.

1.	Meissner A, Mikkelsen TS, Gu H, et al. Genome-scale DNA methylation maps of pluripotent and differentiated cells. *Nature*. Aug 7 2008;454(7205):766-70. doi:10.1038/nature07107

**Supplementary Table S1**

**Study capsule fatty acid composition^a^**

| **Fatty acid^b^** | **5g/d** | **1g/d** |
| --- | --- | --- |
| **6:0** |  | 0.001 |
| **8:00** |  | 0.018 |
| **10:00** |  | 0.014 |
| **12:00** |  | 0.112 |
| **14:00** |  | 0.046 |
| **16:00** |  | 0.139 |
| **16:1n7** |  | 0.005 |
| **18:00** |  | 0.058 |
| **18:1n-9** |  | 0.256 |
| **18:2n-6** |  | 0.401 |
| **18:3n-6** |  | 0.045 |
| **18:3n-3** |  | 0.045 |
| **20:00** |  | 0.003 |
| **20:1n-11** |  | 0.003 |
| **22:00** |  | 0.002 |
| **20:5n-3** | 0.746 | 0.134 |
| **22:6n-3** | 0.318 | 0.049 |
| **Other** | 0.176 | 0.045 |

^a^Nutrition Data System for Research software, version 2019 (Nutrition Data Coordinating Center, University of Minnesota, Minneapolis, MN) used for fatty acid composition, with the exception of EPA, DHA, and other unspecified fish oil FAs which are reported as mean values of two batches based on the Certificates of Authentication (Marine Ingredients).

^b^gram per capsule

**Supplementary Table S2, erythrocyte membrane fatty acid profiles**

| **FattyAcid**^a^ | **Arm**^b^ | **0 months** | **3 months** | **6 months** | **9 months** | **12 months** |
| --- | --- | --- | --- | --- | --- | --- |
| 14:0 | high | 0.30 (.17) | 0.25 (.13) | 0.23 (.13) | 0.21 (.12) | 0.24 (.12) |
|  | low | 0.30 (.15) | 0.31 (.16) | 0.26 (.14) | 0.25 (.11) | 0.28 (.10) |
| 16:0 | high | 23.65 (1.11) | 24.24 (1.39) | 24.07 (1.44) | 23.96 (1.14) | 23.81 (1.11) |
|  | low | 23.72 (1.16) | 23.95 (1.37) | 23.96 (1.33) | 23.44 (1.15) | 23.38 (1.04) |
| 16:1n7 | high^ | 0.62 (.37) | 0.46 (.29) | 0.46 (.29) | 0.42 (.24) | 0.48 (.26) |
|  | low | 0.60 (.27) | 0.56 (.29) | 0.53 (.25) | 0.51 (.19) | 0.54 (.22) |
| 18:0 | high | 19.15 (1.16) | 19.69 (1.16) | 19.48 (.96) | 19.38 (1.06) | 18.99 (1.23) |
|  | low | 19.44 (1.08) | 19.34 (1.29) | 19.59 (1.21) | 19.37 (.89) | 19..23 (.85) |
| 18:1n-9 | high | 13.48 (1.17) | 13.00 (1.13) | 13.23 (1..20) | 13.31 (1.02) | 13.37 (1.26) |
|  | low | 13.73 (1.26) | 13.51 (1.08) | 13.43 (1.24) | 13.52 (1.07) | 13.84 (.80) |
| 18:2n-6* | high^ | 14.41 (1.84) | 11.91 (1.60) | 12.49 (1.97) | 12.67 (1.49) | 13.13 (2.10) |
|  | low | 13.69 (1.86) | 13.60 (1.81) | 13.26 (1.75) | 13.72 (1.89) | 13.96 (1.87) |
| 18:3n-3 | high | 0.21 (.08) | 0.18 (.11) | 0.18 (.07) | 0.16 (.04) | 0.17 (.06) |
|  | low | 0.24 (.14) | 0.21 (.09) | 0.18 (.06) | 0.17 (.05) | 0.19 (.05) |
| 20:2n-6* | high^ | 0.27 (.07) | .19 (.08) | 0.21 (.09) | 0.21 (.07) | .21 (.08) |
|  | low | 0.24 (.07) | 0.22 (.07) | 0.24 (.10) | 0.26 (.10) | 0.27 (.10) |
| 20:3n-6* | high^ | 1.70 (.50) | 1.00 (.21) | 1.00 (.24) | 1.05 (.27) | 1.04 (.31) |
|  | low^ | 1.69 (.36) | 1.49 (.35) | 1.45 (.36) | 1.45 (.31) | 1.44 (.29) |
| 20:4n-6* | high^ | 16.24 (1.84) | 12.31 (1.65) | 11.65 (2.19) | 11.55 (2.26) | 11.49 (1.88) |
|  | low^ | 16.11 (1.52) | 14.75 (1.94) | 14.73 (1.77) | 15.10 (1.50) | 14.79 (1.87) |
| 20:5n-3* | high^ | 0.49 (.20) | 5.15 (1.47) | 5.24 (1.87) | 5.43 (1.83) | 5.33 (2.07) |
|  | low^ | 0.57 (.22) | 1.68 (.45) | 1.73 (.55) | 1.67 (.53) | 1.65 (.57) |
| 22:4n-6* | high^ | 3.58 (.84) | 1.87 (.38) | 1.38 (.54) | 1.34 (.58) | 1.31 (.44) |
|  | low^ | 3.51 (.79) | 2.62 (.60) | 2.41 (.61) | 2.50 (.62) | 2.48 (.69) |
| 22:5n-3* | high^ | 2.17 (.27) | 4.05 (.64) | 4.32 (.68) | 4.24 (.67) | 4.21 (.84) |
|  | low^ | 2.34 (.24) | 3.21 (.39) | 3.50 (.64) | 3.36 (.46) | 3.25 (.43) |
| 22:5n-6* | high^ | 0.55 (.19) | 0.19 (.09) | 0.13 (.09) | 0.12 (.08) | 0.11 (.09) |
|  | low^ | 0.45 (.17) | 0.28 (.12) | 0.24 (.16) | 0.22 (.15) | 0.21 (.11) |
| 22:6n-3* | high^ | 3.18 (.87) | 5.50 (1.34) | 5.94 (1.03) | 5.92 (1.07) | 6.10 (1.04) |
|  | low^ | 3.39 (1.17) | 4.26 (.91) | 4.49 (1.07) | 4.45 (.98) | 4.49 (.98) |
| Total n-3* | high^ | 6.04 (1.08) | 14.88 (2.61) | 15.67 (3.28) | 15.76 (3.26) | 15.82 (3.60) |
|  | low^ | 6.53 (1.08) | 9.36 (1.40) | 9.90 (1.81) | 9.65 (1.64) | 9.58 (1.67) |
| Total n-6* | high^ | 36.75 (2.09) | 27.47 (2.95) | 26.87 (3.85) | 26.95 (3.58) | 27.29 (3.63) |
|  | low^ | 35.69 (2.09) | 32.96 (2.36) | 32.33 (2.50) | 33.26 (2.04) | 33.15 (2.04) |
| Total MUFA | high | 14.11 (1.27) | 13.46 (1.18) | 13.69 (1.30) | 13.73 (1.07) | 13.85 (1.42) |
|  | low | 14.33 (1.23) | 14.07 (1.07) | 13.96 (1.28) | 14.03 (1.09) | 14.38 (.78) |
| Total SFA | high | 43.10 (1.17) | 44.19 (1.83) | 43.78 (1.54) | 43.56 (1.46) | 43.05 (1.28) |
|  | low | 43.45 (1.61) | 43.60 (1.79) | 43.81 (2.08) | 43.06(1.28) | 42.89 (1.06) |
| n-6:n-3* | high^ | 6.31 (1.31) | 1.95 (.66) | 1.88 (.90) | 1.87 (.88) | 1.91 (.86) |
|  | low^ | 5.74 (1.37) | 3.63 (.82) | 3.44 (1.09) | 3.58 (.90) | 3.61 (.92) |
| O3FAI^c^ | high | 3.66 (0.98) | 10.65 (2.21) | 11.18 (2.72) | 11.36 (2.66) | 11.43 (2.91) |
|  | low | 3.96 (1.34) | 5.94 (1.18) | 6.22 (1.49) | 6.12 (1.37) | 6.14 (1.43) |

^a^Fatty acids as percentage of total fatty acids (weight %) with standard deviation (SD)

^b^*n* subjects at 0, 3, 6, 9, 12 mos = 32, 27, 27, 25, 25, respectively, in high dose and 32, 28, 27, 24, 25, respectively, in low dose arms.

^c^omega-3 index = EPA+DHA content of erythrocytes as percentage total fatty acids (weight %)

*p-values<0.001, 0-12 months between arms except 18:2n-6=.032 by MMRM

^p-values<0.001, 0-12 months within arm except 18:2n-6=.008, 16:1n7=.024

**Supplementary Figure S1:**

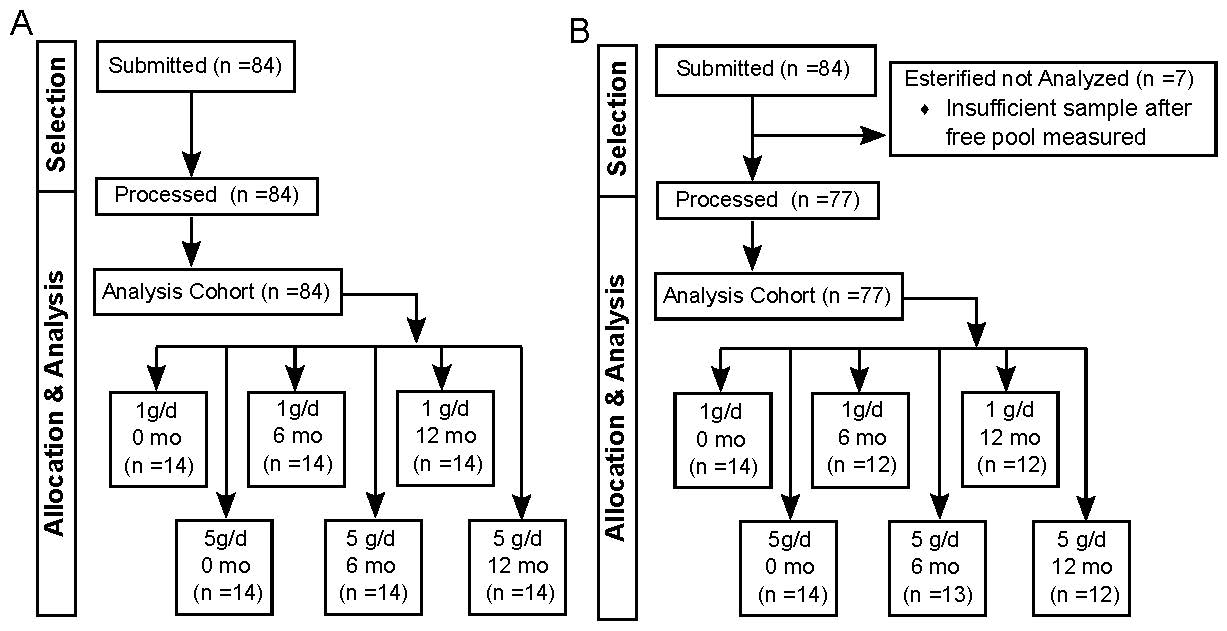

**Supplemental Figure S1: Oxylipin analyses cohort diagram.** A total of 84 samples were submitted for analysis. A) All samples were processed for non-esterified oxylipins. B) Seven samples had insufficient material for the additional analysis of esterified oxylipins.

**Supplementary Table S3**

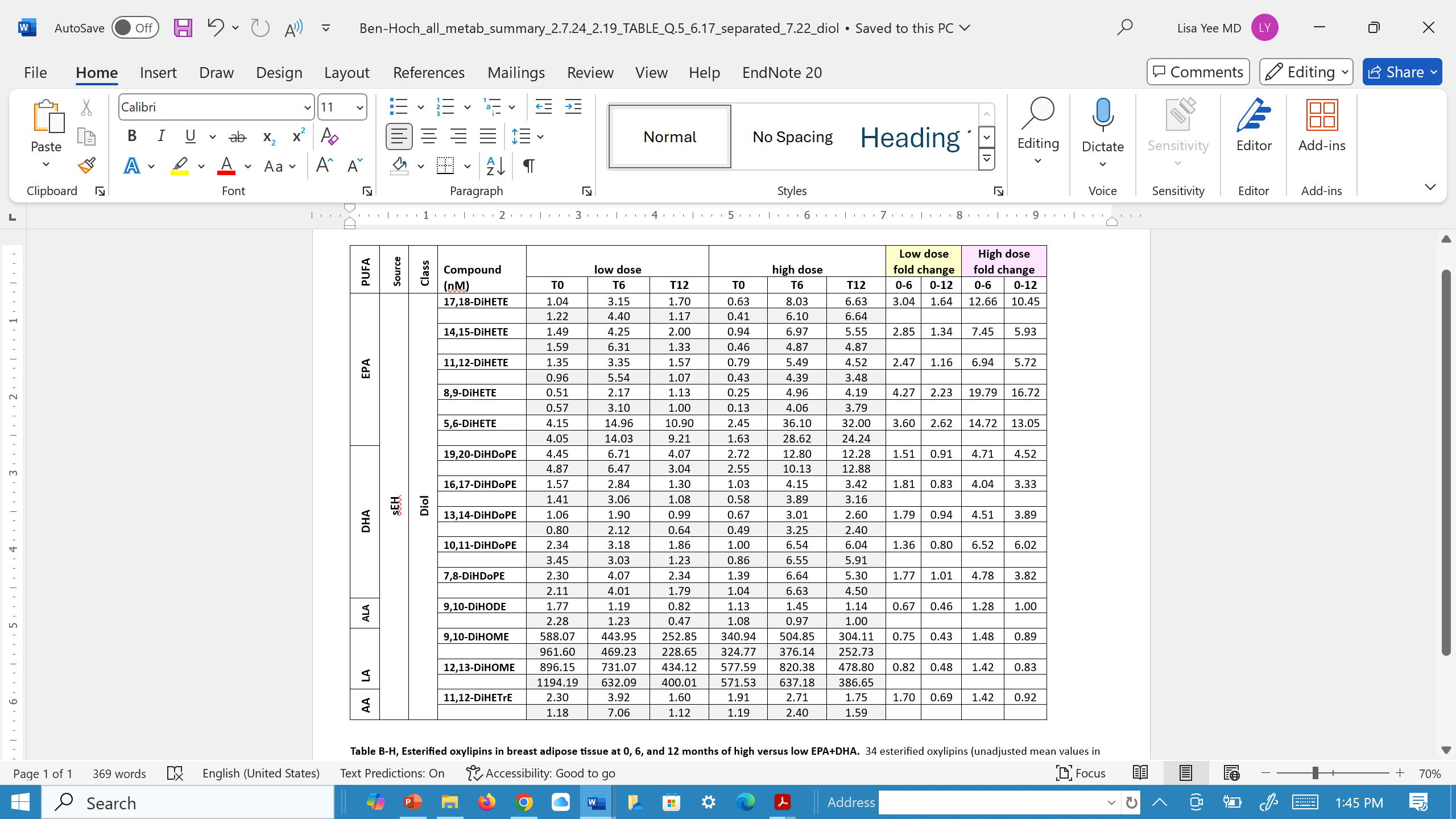

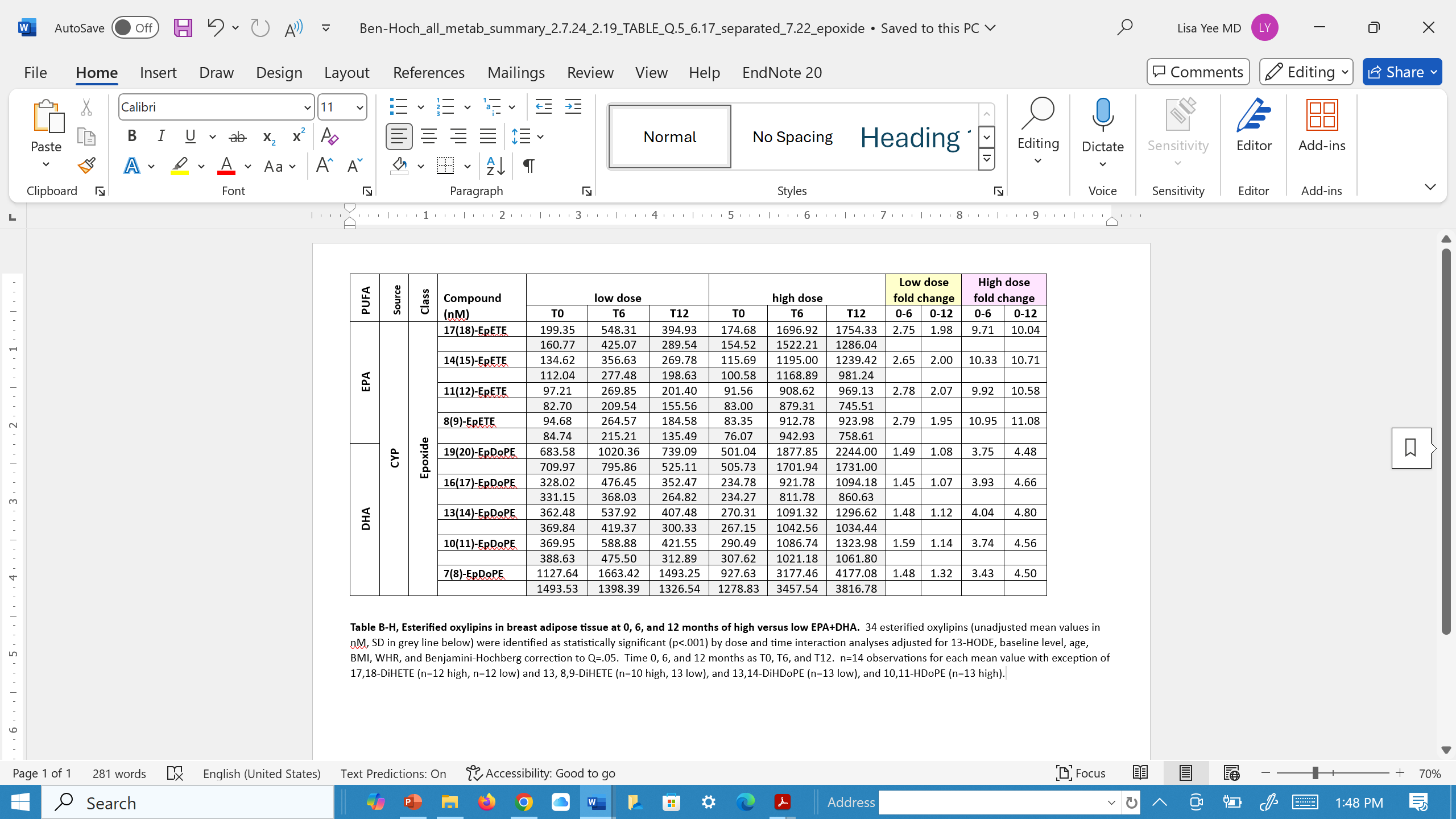

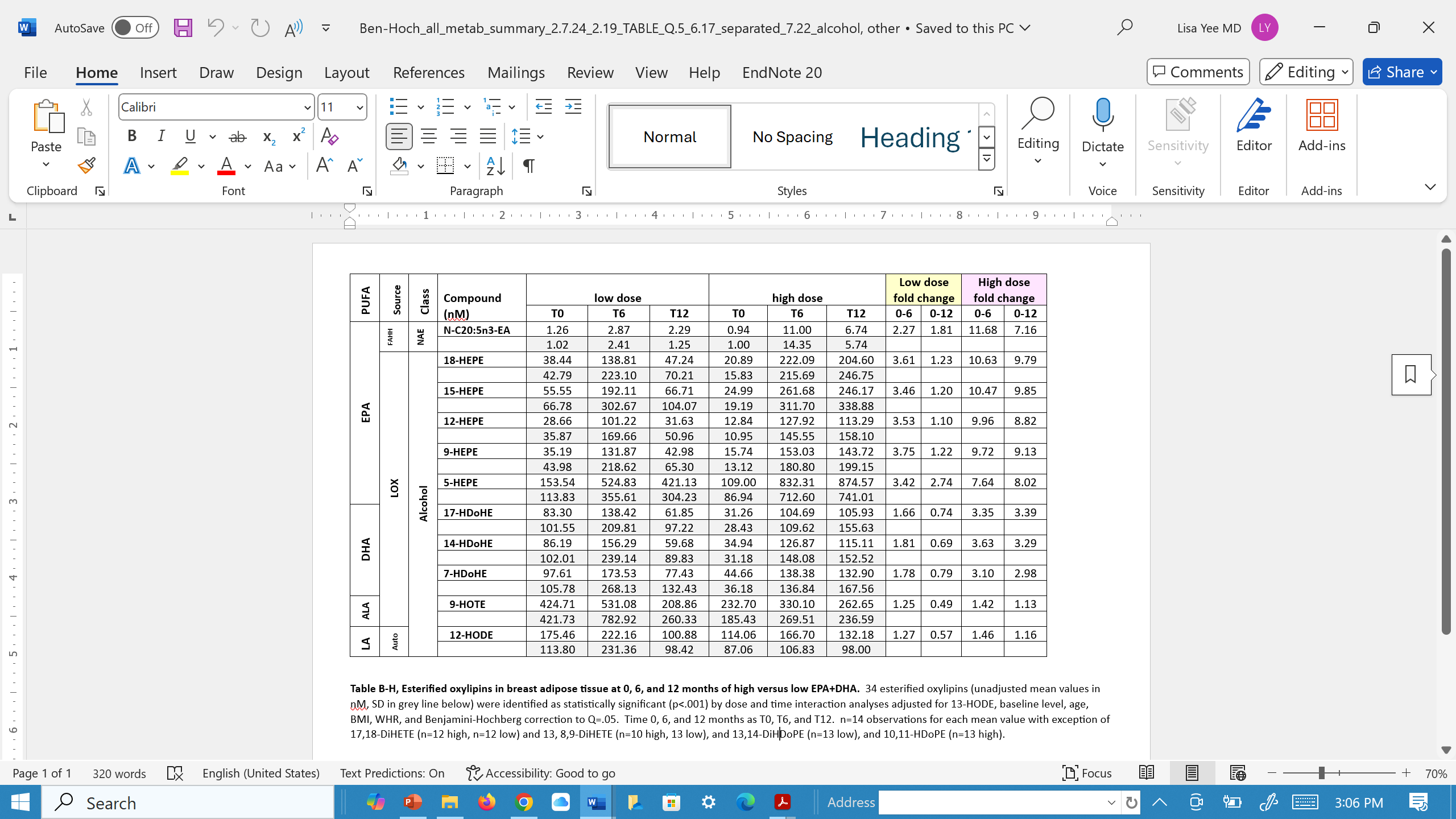

**Supplementary Table S3, Esterified oxylipins in breast adipose tissue at 0, 6, and 12 months of 5g/d vs 1g/d EPA+DHA.** 34 esterified oxylipins (unadjusted mean values in nM, SD in grey line below) were identified as statistically significant (p<.001) by dose and time interaction analyses adjusted for 13-HODE, baseline level, age, BMI, WHR, and Benjamini-Hochberg correction to Q=.05. Time 0, 6, and 12 months as T0, T6, and T12. n=14 observations for each mean value with exception of 17,18-DiHETE (n=12 high, n=12 low) and 13, 8,9-DiHETE (n=10 high, 13 low), and 13,14-DiHDoPE (n=13 low), and 10,11-HDoPE (n=13 high).

**Supplemental Table 4.1: DMR enrichment in inflammation pathways**

Number of DMRs found in each inflammation-related pathway. Bionomial p-value = 1 for all comparisons.

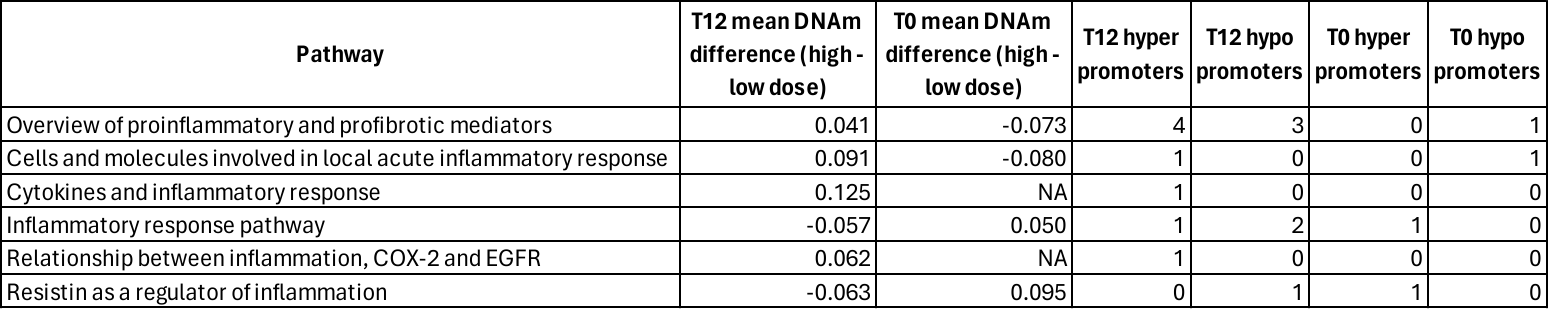

**Supplemental Table 4.2: Inflammation-related DNAm changes.**

The gene promoters from inflammation pathways that were found to differentially methylated between high and low dose arms are shown.

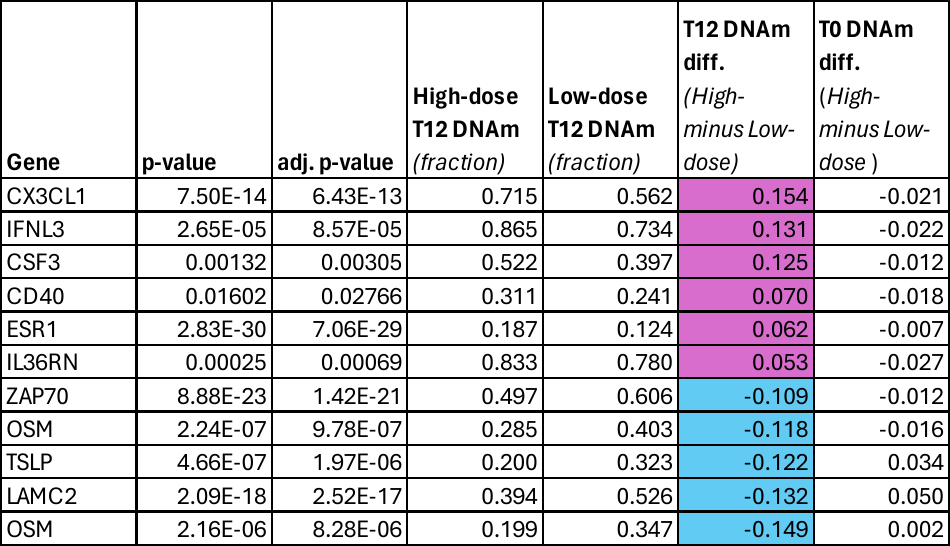

**Table S4.3: Metabolism pathway DNAm analysis**

All metabolism-related pathways that were tested for concerted DNAm changes between the high and low dose arms at the 12-month timepoint.

| **Pathway** | **FDR**  **p-value 12 mos** | **FDR**  **p-value 0 mos** | **Mean DMR DNAm 12 mos** | **Mean DMR DNAm 0 mos** | **Hyper DMRs 12 mos** | **Hypo DMRs 12 mos** | **Hyper DMRs 0 mos** | **Hypo DMRs 0 mos** |
| --- | --- | --- | --- | --- | --- | --- | --- | --- |
| Orexin receptor pathway | 0.031 | 1 | 0.068 | -0.068 | 14 | 2 | 0 | 3 |
| SREBP signaling | 0.031 | 1 | 0.088 | -0.072 | 8 | 0 | 0 | 3 |
| Eicosanoid metabolism, COX | 0.063 | 1 | 0.131 | NA | 6 | 0 | 0 | 0 |
| Leptin signaling pathway | 0.063 | 1 | 0.062 | -0.061 | 6 | 0 | 0 | 1 |
| Regulation lipid metabolism, PPARα | 0.200 | 1 | 0.079 | NA | 4 | 0 | 0 | 0 |
| Proinflammatory, profibrotic mediators | 0.677 | 1 | 0.041 | -0.073 | 6 | 3 | 0 | 1 |
| PI3K-Akt pathway | 1.000 | 1 | -0.001 | -0.021 | 5 | 4 | 1 | 2 |
| Ras signaling | 1.000 | 1 | -0.026 | 0.056 | 2 | 2 | 1 | 0 |

**Figure S2. DNAm change in the Orexin pathway**

DNAm changes in the orexin pathway indicate a dominant hypermethylation pattern in breast adipose tissue following 12 months of 5g/d EPA+DHA.

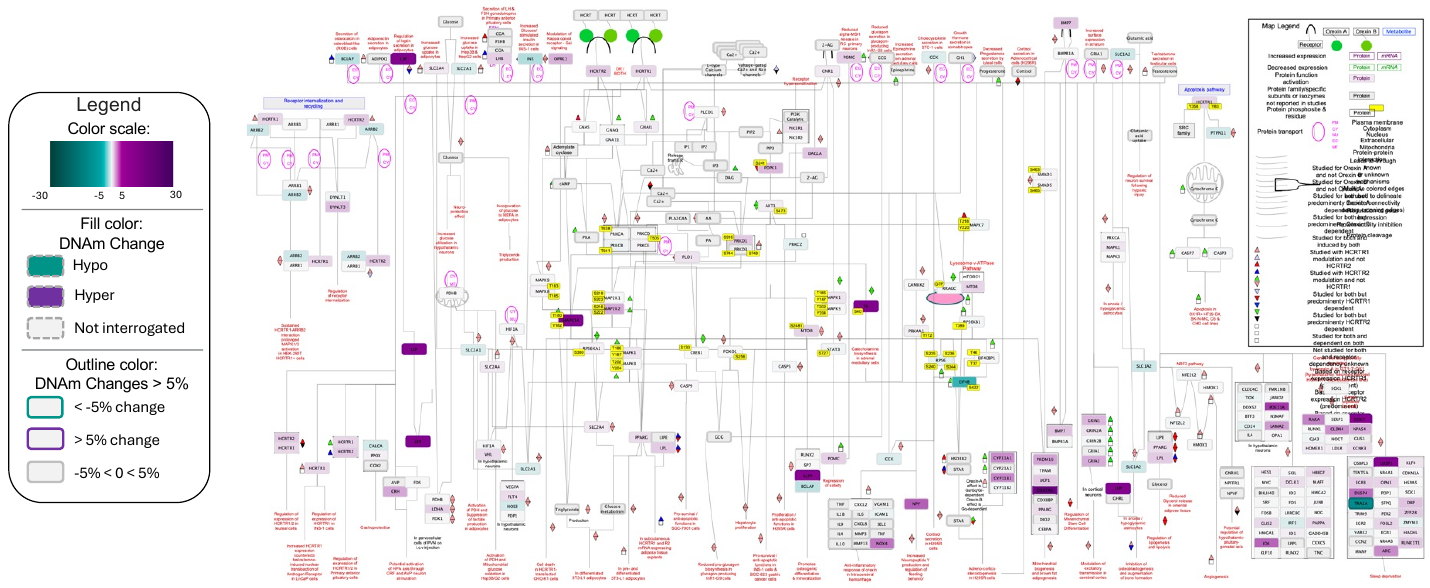
